# Inferring SARS-CoV-2 RNA shedding into wastewater relative to time of infection

**DOI:** 10.1101/2021.06.03.21258238

**Authors:** Sean Cavany, Aaron Bivins, Zhenyu Wu, Devin North, Kyle Bibby, T. Alex Perkins

## Abstract

Since the start of the COVID-19 pandemic, there has been interest in using wastewater monitoring as an approach for disease surveillance. A significant uncertainty that would improve interpretation of wastewater monitoring data is the intensity and timing with which individuals shed RNA from severe acute respiratory syndrome coronavirus 2 (SARS-CoV-2) into wastewater. By combining wastewater and case surveillance data sets from a university campus during a period of heightened surveillance, we inferred that individual shedding of RNA into wastewater peaks on average six days (50% uncertainty interval (UI): 6 – 7; 95% UI: 4 – 8) following infection, and that wastewater measurements are highly overdispersed (negative binomial dispersion parameter, *k* = 0.39 (95% credible interval: 0.32 – 0.48)). This limits the utility of wastewater surveillance as a leading indicator of secular trends in SARS-CoV-2 transmission during an epidemic, and implies that it could be most useful as an early warning of rising transmission in areas where transmission is low or clinical testing is delayed or of limited capacity.

---

Since the onset of the COVID-19 pandemic, there have been over 170 million known SARS-CoV-2 infections^1^. From early on in the pandemic, reporting delays and changes in testing effort and capacity have made timely surveillance difficult^2–5^. This led to interest in using the concentration of SARS-CoV-2 RNA in wastewater as a tool for COVID-19 surveillance and to monitor for secular trends. In the past, this type of surveillance has been used to provide early warnings of polio outbreaks^6^ and to monitor for antimicrobial-resistant pathogens^7^. Initial hope that wastewater could be used as a leading indicator of SARS-CoV-2 transmission has led to mixed results^8,9^.

Understanding how temporal patterns in the incidence of new infections relates to the observed concentration of SARS-CoV-2 RNA in wastewater is key to interpreting wastewater surveillance data. These two quantities can be linked by the distribution, relative to the time of infection, of individual shedding of viral RNA into the wastewater system. This is analogous to the way in which the incidence of infection is linked to the timing of symptom onset by the incubation period in epidemiology. SARS-CoV-2 RNA has been observed in stool samples as early as a few days after hospital admission^10^ and within a week of symptom onset^11^, and as late as five weeks after respiratory samples are no longer positive for SARS-CoV-2 RNA^12^. The intensity with which SARS-CoV-2 is shed across this wide range of times relative to infection is presently unclear.

We used data from a COVID-19 outbreak on a university campus^13^ to infer the shedding distribution of SARS-CoV-2 in wastewater relative to time of infection, combining daily COVID-19 case notifications and daily measurements of SARS-CoV-2 RNA in the university sewage system. As approximately 85% of students live on campus ^14^, the sewage system is largely representative of the student body as a whole, which, when coupled with intense on-campus case surveillance and an outbreak with two distinct temporal peaks, makes this an ideal dataset for estimating the shedding distribution.

The majority of students arrived on campus in the week August 3 – 9 for the start of classes on August 10. Soon after students arrived, there was a large number of reported COVID-19 cases among students, peaking at 177 cases reported on August 17 (Fig. 1A). In response to this, the University underwent a short period of online instruction from August 19 to September 2, while students remained on campus. Case notifications declined thereafter and remained at a lower level, until rising again in October and November. In total, there were 2,263 cases among students between the start and end of the semester (November 20). In the earlier part of the semester, the majority of notified cases were symptomatic when notified, though some asymptomatic and pre-symptomatic cases were detected through contact tracing and limited surveillance testing in specific groups, such as athletes. During the semester, surveillance testing capacity was substantially increased, and hence later in the semester the majority of cases notified were not symptomatic at the time of detection (Fig. 1A). Throughout the semester, a proportion of students were isolated off campus for 10 days following a positive test result (Fig. S1). This proportion declined slightly over the semester, with about 25% of cases isolated off campus over the entire semester.

**Fig 1.**
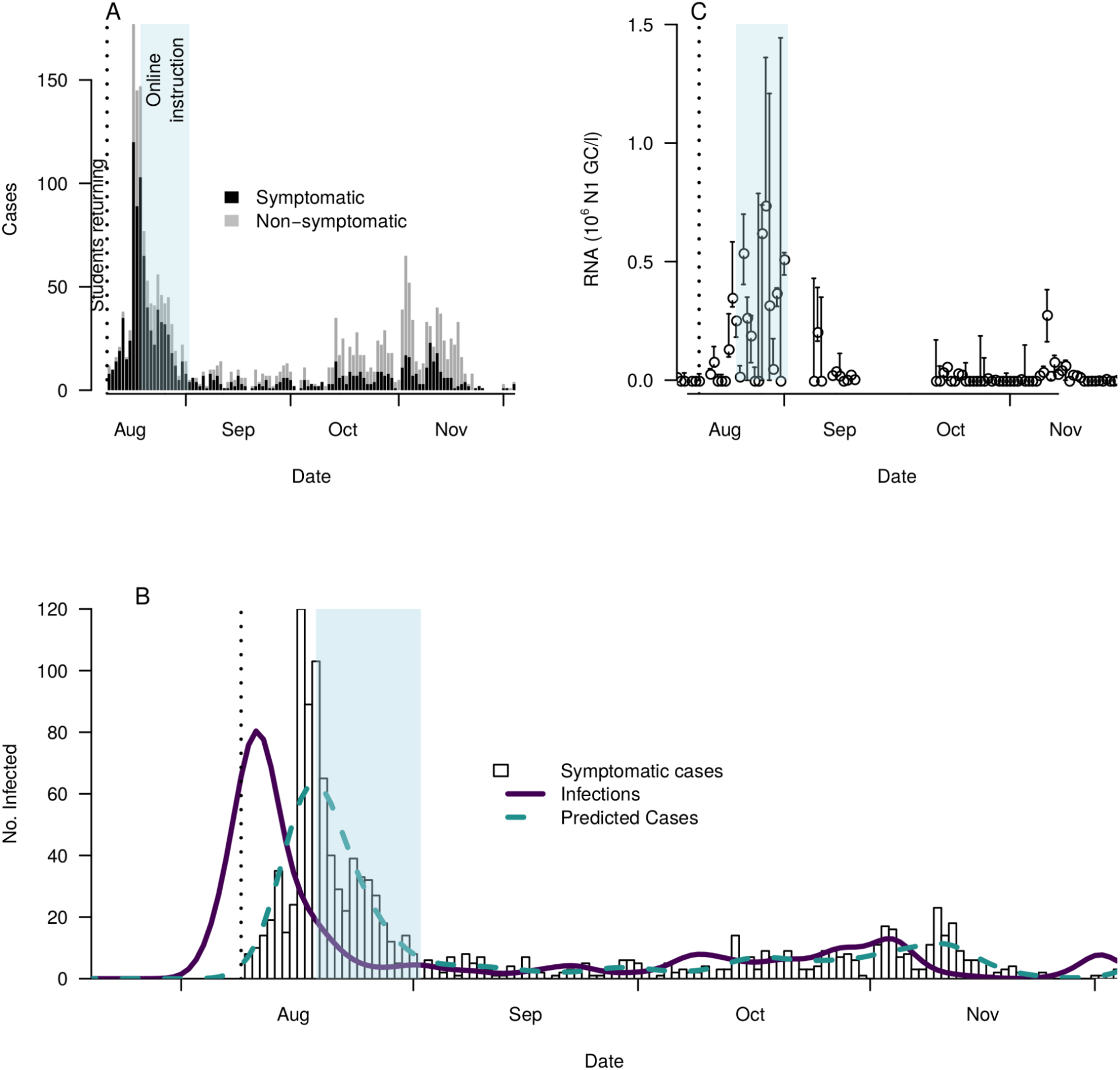
In all panels the shaded blue region indicates a period of online instruction, where students remained on campus but did not have in person instruction, and the vertical dashed line indicates the start of classes (August 9); the majority of students arrived in the week preceding this date. A: notified cases among students on the University campus in the fall semester, delineated by whether the case was found due to symptomatic screening or not. B: the estimated timeseries of infections (purple line), alongside the actual (bars) and implied (green dashed line) distributions of symptomatic case notification dates. C: wastewater measurements by date. Open circles indicate the daily mean of three recovery-corrected RNA measurements, and whiskers indicate the smallest and largest measurements on that day.

From the time series of notified cases, we estimated the time series of infection incidence by deconvolving the case notifications with the distribution of the time between infection and testing using a maximum-likelihood approach. Due to the increase in surveillance testing over the course of the semester, we limited this to cases that were found through symptomatic testing. For these cases, we modeled the time from infection to test result as the incubation period plus the delay between symptom onset and test result. We assumed this delay was Poisson distributed with mean 2 days, and explored other values of the delay in a sensitivity analysis (Methods). When we used this infection time-series to project back the case-notification time series from which it was derived, we were able to recover the timing of peaks in notifications, although the height of the initial peak was underestimated. We estimate that a bare minority of transmission relating to the first large peak occurred prior to classes starting, as students were arriving back on campus, and that transmission was already rapidly declining when online classes began (Fig. 1B). Pre-arrival testing of all students ascertained 33 cases out of 11,836 tests.

Throughout this period, we collected one 24-hour time-based composite sample per day from a manhole collecting all the wastewater produced on campus and evaluated the concentration of SARS-CoV-2 RNA in these samples (Fig. 1C). Sewage samples were concentrated using electronegative membrane filtration, homogenized, and extracted using a commercial kit. SARS-CoV-2 RNA in each sample was enumerated using reverse transcription droplet digital PCR (RT-ddPCR) performed in triplicate resulting in three measurements of SARS-CoV-2 RNA in wastewater per day (Methods). The recovery efficiency of the sampling process was assessed by adding an exogenous control virus at a known concentration to wastewater samples. SARS-CoV-2 RNA concentrations in wastewater were recovery-corrected to account for the measured process efficiency. As expected, the RNA concentration showed a similar temporal pattern to cases, though substantially more noisy. The cross-correlation function of RNA concentration and case notifications was highest at a lag of 7-9 days and was not significant at the 95% level at lags less than -1 days, implying that the wastewater was likely not a leading indicator of trends in case numbers in this context of relatively rapid case notifications (Fig. S2).

To estimate the individual shedding distribution, we first assumed the average intensity of individual shedding was gamma distributed through time, given its flexible shape. We then reconstructed the RNA concentration in the wastewater using this assumed average shedding distribution by taking our estimated infection time series and then summing the shedding distributions for each infection through time. We accounted for those in off-campus isolation by reducing the individual shedding distribution according to the fitted probability that they were isolated off-campus (Fig. S1) and the distribution of dates they may have entered and exited quarantine, the latter estimated from the distribution of the time between infection and a positive test result. We then scaled the magnitude of the shedding distribution by a constant to capture the magnitude of shedding intensity. We calibrated the parameters of the gamma distribution and the peak shedding intensity to the sewage data using Markov chain Monte Carlo methods, with a negative binomial likelihood to allow for overdispersion in sewage measurements (Methods). We also calibrated a fixed level of underreporting, with unreported infections assumed to not enter quarantine or isolation. See Fig. 3 for examples of how different gamma distributions and delays from symptom onset to testing translate into different temporal patterns of viral RNA measurements in wastewater.

The shedding distribution we inferred rises rapidly following infection and through the incubation period, peaking at around 6 days (50% uncertainty interval (UI): 6 – 7; 95% UI: 4 – 8) following infection (Fig. 2A, S4A). The estimated genome concentration in a given sample was highly overdispersed, with only 28% of predicted samples being above the 95% limit of detection at the predicted peak SARS-CoV-2 RNA concentration (Fig. 2B, S4B). This is likely a combination of overdispersion in individual shedding and overdispersion from sampling due to the sewage not being well mixed at the point of sample collection. The maximum *a posteriori* estimate of the dispersion parameter was 0.393 (95% Credible Interval: 0.317 – 0.478).

**Fig 2.**
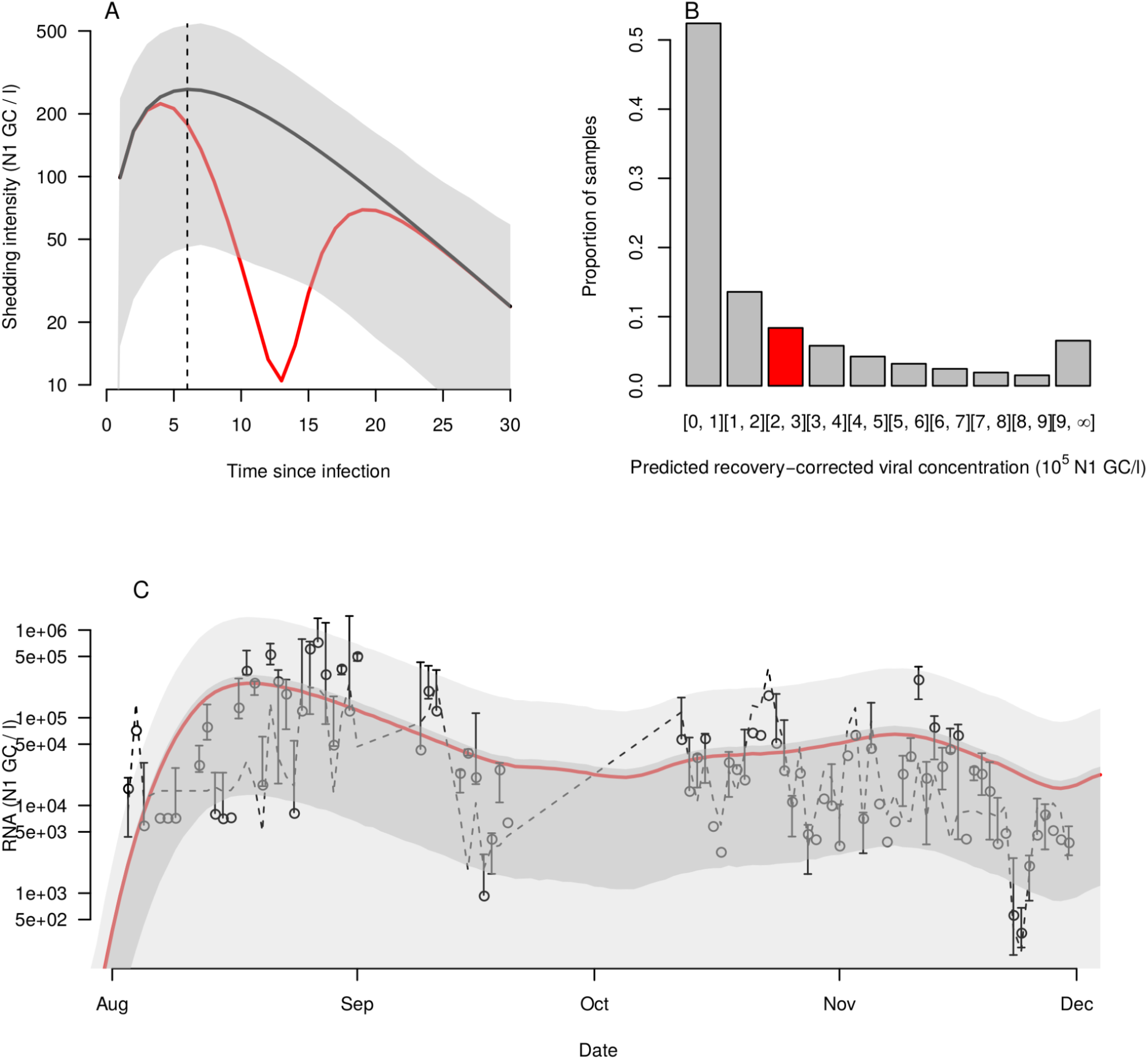
A: Implied shedding distribution and 95% credible interval. The vertical dashed line indicates the day of peak shedding (eight days). The red line indicates the average shedding distribution of someone who enters isolation B: Distribution of predicted daily recovery-corrected measurements at the peak intensity (August 16) implied by the negative binomial likelihood. The red shaded bar shows the range in which the predicted mean measurement fell C: Mean predicted RNA concentration (red line), 95% prediction interval (light gray line). White open circles and whiskers indicate the daily mean measurements and minimum/maximum values respectively. The dashed line indicates the recovery-corrected 95% limit of detection. Data points below the 95% limit of detection are plotted as the midpoint of 0 and the 95% limit of detection for that day.

To evaluate the fit of our inferred shedding distribution, we reconstructed the predicted concentration of SARS-CoV-2 in the wastewater (Fig. 2C, S4C). The mean predicted concentration captures the trends in viral concentration, though it peaks slightly too early—the peak in the mean predicted concentration occurred on August 18, while the highest data point was recorded on August 31 and the highest daily mean and median on August 27. The predicted time series were also very noisy, implying that it is necessary to average multiple daily samples to gain an impression of temporal trends, particularly due to variability in composite sampling or when the number of infections is small. The upper bound of our 95% prediction interval (i.e., the 97.5^th^ percentile) was greater than 96.6% of measured values, and the upper bound of our 50% prediction interval (i.e., the 75^th^ percentile) was greater than 81.6% of measured values. The Spearman’s rank correlation coefficient of the mean predicted concentration and the mean daily estimated concentration was 0.51 (95% prediction interval: -0.01 – 0.54). When we increased or decreased the mean delay from symptom onset to testing, the predicted peak in the shedding distribution was a similar amount of time later or earlier respectively, but the overall shape of the distribution remained the same, and in particular its long tail (Figs S5-7).

The shedding distribution that we inferred peaks on average around 6 days after infection, before the onset of symptoms in 35% (95% credible interval (CrI): 15% - 73%) of patients that become symptomatic ^15^. Following this peak, the shedding intensity decays approximately exponentially, though at a lower rate than the initial increase. This slow decay means that, on average 77% (95% CrI: 44% - 95%) of shedding occurs after the end of the incubation period This early peak and slower exponential decay is broadly in line with measurements taken from stool samples by Wölfel et al.^11^ and a meta-analysis of shedding in stool by Cevik et al.^16^. The early peak and slower decline indicates one reason why it is difficult to use wastewater data as a leading indicator: while some recent infections will be shedding, that signal may be masked by a large number of older infections. This is particularly the case in a period of declining or stable transmission, when the day-6 peak and the long tail in shedding suggest that much of the RNA measured may come from people infected some time ago. For this reason, wastewater surveillance may be most useful to detect increased transmission in an area where there has recently been little or no transmission, or where there are greater testing delays and/or limited testing capacity^17^. One can observe this early warning in data from the end of July in our data set, where positive wastewater samples were recorded in advance of an increase in cases (Fig. 1A and C). If incidence stabilizes at a relatively high level following an initial outbreak, the long tail in the shedding distribution implies that we may counterintuitively continue to see rising RNA concentration measurements for several weeks after case numbers have declined (see Fig. S8 for examplar incidence curves and the associated predicted mean wastewater measurements).

A modeling study by Huisman et al., whose aim was to estimate the reproduction number over time (*R*_*t*_) directly from wastewater data, also estimated which shedding distribution minimized the error between estimates of *R*_*t*_ based on wastewater and that based on cases^18^. Their inferred distribution had a much sharper peak, but also peaked around 6 days following infection. However, that study used data from larger populations and catchment areas and had a different and less direct calibration procedure, which may explain the discrepancy in shedding distribution shape. Another study (Schmitz et al. ^19^) attempted to estimate individual shedding using data from a different university campus, finding that the mean shedding rate was 6.84 log_10_ N1 GC per gram of feces. However, it is difficult to compare this directly to our estimates, as the Schmitz et al. study does not have a temporal component and is per gram of feces, where ours is per liter of wastewater and agnostic to shedding route.

Our study has several limitations. First, we assumed that the distribution of testing delays was the same through time. In reality, there may have been longer delays earlier in the semester when testing capacity was lower and the number of cases was higher. Second, underreporting of cases means that our infection timeseries may not reflect the true magnitude of infections. If underreporting was constant through time, then we would still capture the temporal patterns in new infections, and hence the temporal distribution of shedding intensity would be correctly inferred, though if under-reporting increased over time this would lead to incorrect inference. We estimated the proportion of cases that were detected through symptomatic testing, assuming this was constant through time, to be 0.57 (95% CrI: 0.11 - 0.98). The high degree of uncertainty on this parameter estimate leads to great uncertainty in the magnitude of individual shedding. Third, our population was predominantly students, most of whom were between the ages of 18 and 22, and so extrapolations to other age-groups may not be appropriate. Finally, our inferred shedding distribution reflects the contribution each infected individual makes per liter of sampled wastewater, rather than per liter of wastewater produced by that individual. It therefore includes the effect of dilution. While this makes it difficult to estimate the magnitude of individual shedding, it should not affect our estimate of its temporal distribution.

In summary, we have estimated that infected individuals likely shed SARS-CoV-2 RNA into wastewater for a prolonged period, peaking at around 6 days after infection, longer than the incubation period for COVID-19. This implies that wastewater data may be most useful to detect new outbreaks when incidence is low, when testing capacity is low, or when test results are substantially delayed. It also highlights that care must be taken when interpreting wastewater data during an ongoing period of high incidence.

## Methods

### Campus data

Approximately 7,000 undergraduate students live on the University campus. Early in the semester, the majority of cases were notified through screening of individuals with symptoms (Fig. 1A). These individuals were tested using RT-PCR. Following the initial outbreak, surveillance screening increased, resulting in more individuals being diagnosed who were either pre- or asymptomatic. Screening of non-symptomatic individuals was done with rapid saliva antigen tests, with individuals positive on this test being followed up with confirmatory RT-PCR tests before being notified as a case. Students arrived back on campus in time for the start of classes on August 9; most students arrived back in the final week before classes started, i.e. beginning on August 2. Following high incidence in the first weeks back, the University underwent a two-week period of online classes, starting on August 19 and ending on September 2. Throughout the semester, all students were required to wear masks at all times in public places and to socially distance, while class sizes were reduced. Following notification, cases that lived on campus entered an isolation period, which typically lasted 10 days. While some of these cases were isolated in on-campus facilities, some cases were isolated off-campus, meaning they would not have been shedding into the campus wastewater system for that period. Additionally, on-campus contacts of notified cases were required to enter a period of quarantine of variable length dependent on test results^20^.

### Wastewater data

From August 3 to November 30, 2020, 24-hour time-based composite wastewater samples were collected each day from the manhole receiving all of the University wastewater. Sampling was interrupted from September 1 to September 9 due to a breakdown of the composite sampler. Full procedures for the processing of the wastewater samples has been detailed elsewhere^21^. Briefly, the wastewater composite sample from each day was mixed well and a 100 mL aliquot was removed, spiked with a process control, and filtered using an electronegative membrane. After filtration, the membranes were aseptically rolled into bead tubes and homogenized using a bead beater, and the resulting liquid was extracted using a Qiagen PowerViral DNA/RNA kit. As detailed elsewhere ^22^, the resulting purified nucleic acids were assayed in triplicate for SARS-CoV-2 RNA using the N1 assay and for the process control RNA in single reactions via RT-ddPCR. During the period of September 21 to October 11, Qiagen PowerViral DNA/RNA kits were not available and an alternative extraction method was used; however, the resulting RT-ddPCR data from this interval did not pass quality assurance protocols and were removed from the resulting dataset.

### Infection timing estimation

To reconstruct the incidence of new infections, we deconvolved the symptomatic case notification time series with the distribution of the time from symptom onset to testing. We modeled the time from infection to testing as the convolution of the incubation period and a delay from symptom onset to testing. We modeled the incubation period as a log-normal distribution with parameters *μ* = 1. 621 and *σ* = 0.418, according to Lauer et al.^15^, and the delay between symptom onset and testing as a Poisson distribution. In the baseline scenario, we used a mean delay of 2 days, but also tried 0, 1, and 5 days in sensitivity analyses (Fig. S5-7). We then deconvolved this distribution from infection to testing from the case notification time series. We first tried performing this deconvolution using a fast Fourier transform, but this approach led to many negative values for infections. Instead, we used maximum-likelihood deconvolution, using the backprojNP function in the R ‘surveillance’ package version 1.18.0^23–25^. This algorithm uses a Poisson likelihood for the number of infections in a day, but makes no assumption on the temporal pattern of infections. We set the smoothing parameter *k* = 10 and used default settings for all other parameters.

### Shedding distribution inference

We modeled the individual shedding distribution, *σ*(*t*), as a gamma distribution with shape *α* and rate *β*, with the origin being the date of infection for that individual. We then adjusted this for entry and exit from isolation using the probability that the individual had entered isolation *t* days after infection, *p*_*enter*_(*t*), the probability they had exited isolation by *t* days after infection, *p*_*exit*_(*t*), the probability that they are reported, *p*_*r*_, and the proportion of reported cases entering isolation on day *s* of the epidemic, *p*_*i*_(*s*). The parameter *p*_*enter*_(*t*) was given by the cumulative probability that the incubation period and delay from symptom onset to testing (described in the previous section) had been completed by day *t*, and *p*_*exit*_(*t*) was just this lagged by 10 days—i.e.

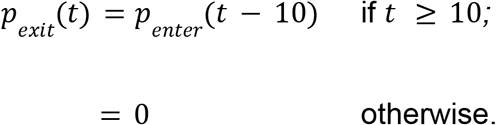

We estimated *p*_*i*_(*s*) by fitting a generalized additive model with a logit link to the proportion of infections entering isolation on day *s*, using the mgcv package (version 1.8-31) in R^26^ (Fig. S1). We estimated *p*_*r*_ in the calibration (see below), assuming that it did not vary through time. Given all these parameters, the individual shedding distribution adjusted for isolation was

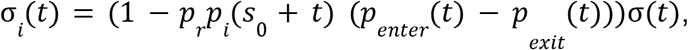

where *s*_0_ is the day of the epidemic on which the individual was infected. Note that as

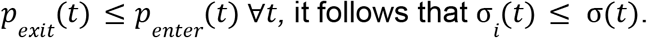

The number of new infections per day, *I*(*t*), was found by dividing the number of infected individuals on a given day that were ultimately reported, *I’*(*t*), by the proportion of infections that were reported, such that 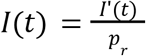. We then estimated the expected temporal pattern of SARS-CoV-2 RNA in the wastewater over time by summing the shedding distribution for each individual infected on a given day through time. We then multiplied this by a scaling constant, *θ*, to capture the magnitude of shedding. This yielded the reconstructed mean RNA concentration,

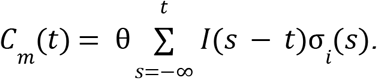

We fitted *C*_*m*_(*t*) to the observed RNA concentration in wastewater, *C*_*d*_(*t*), using Markov chain Monte Carlo methods in the BayesianTools (version 0.1.7) R package^27^, with a negative binomial likelihood, for which the dispersion parameter, *r*, was also fitted. When the RNA concentration in the wastewater was below the 95% limit of detection, we use the cumulative distribution function of the negative binomial distribution to determine the probability of observing data below that threshold. The baseline 95% limit of detection was 220 GC / l, which was then recovery-corrected, *D*_95_ (*t*), and so varies through time (Fig. 2C). As a result, there were five free parameters: *α, β, θ, r*, and *p*_*r*_, with their likelihood given by

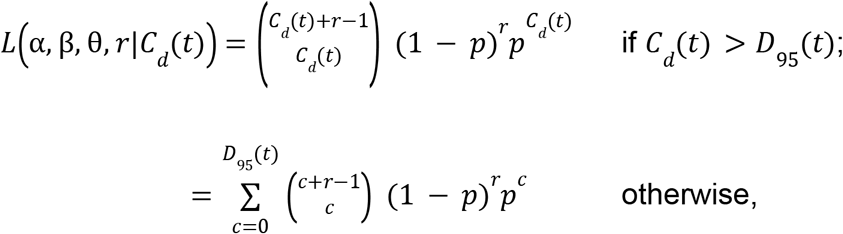

where 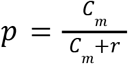. We used default settings from the BayesianTools package, and a uniform prior on all parameters with the following ranges: α ∈ (1, 10^3^), β ∈ (0, 10^3^), θ ∈ (0, 10^5^), *r* ∈ (0, 10^5^), and *p*_*r*_ ∈ (0, 1). We insisted that the shape parameter *α* was greater than 1 so that *σ*(0) *=* 0; i.e., people are not shedding at the moment they are infected. The MCMC chains and posterior parameter densities are shown in Fig. S9 and correlations between parameters are shown in Fig. S10. There were strong positive correlations between *α* and *β* and between *θ* and *p*_*r*_. In the former case, this was likely due to the fact that the mean of the gamma distribution was *α/β*. In the latter case, this was likely due to the fact that *θ* and *p*_*r*_ appear as a ratio in the equation for *C*_*m*_*(t)*, although *p*_*r*_ does also appear in the isolation adjustment of the shedding distribution, so the two parameters were identifiable.

## Data Availability

All underlying data and code to fit model and produce plots is available at https://github.com/scavany/wastewater_shedding_period

## Acknowledgements

This work was funded in part by NSF RAPID grant 2027752. We would like to thank the Notre Dame COVID response team for their work.

## Supplementary figures

**Fig. S1:**
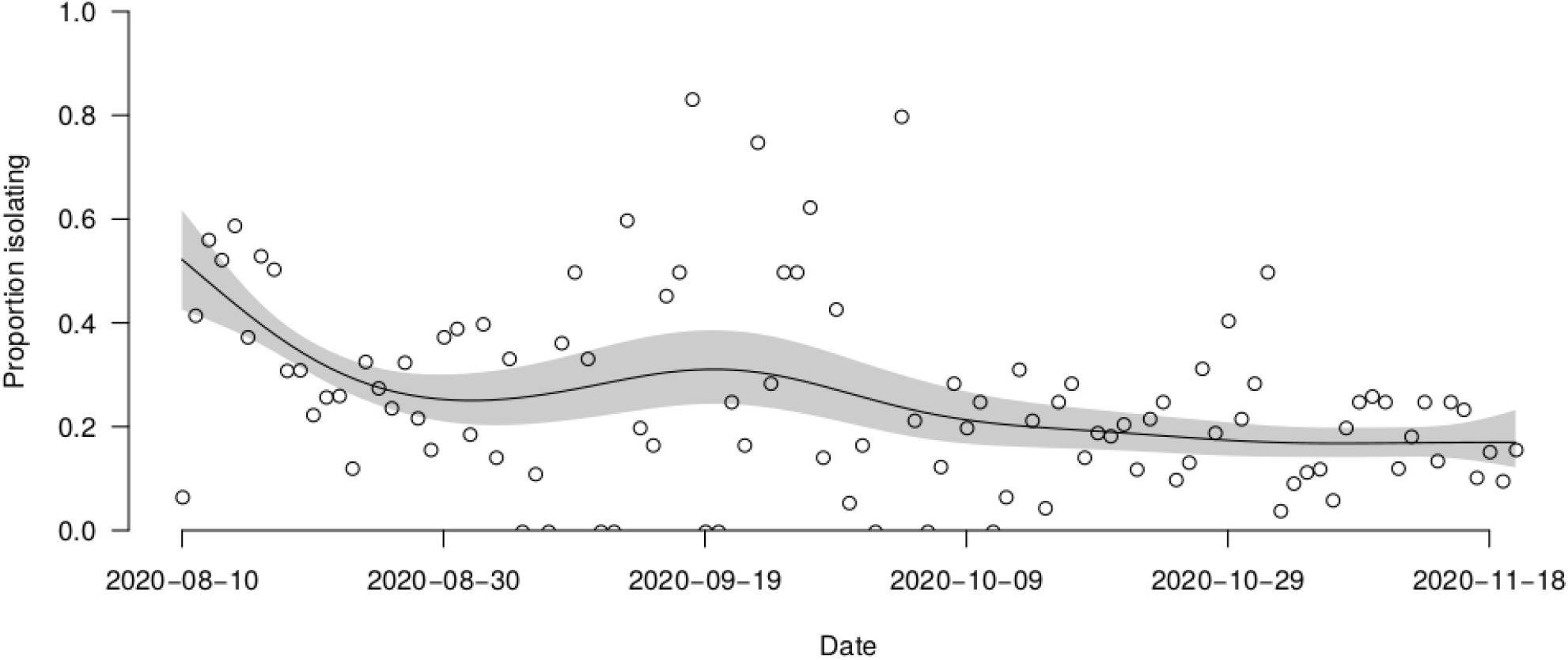
proportion of cases isolating in off-campus housing. Open circles indicate the proportion of notified cases that day that entered isolation that day. Black line and shaded region shows a generalized additive model with logit link fitted to this data, and the 95% confidence interval, respectively.

**Fig. S2:**
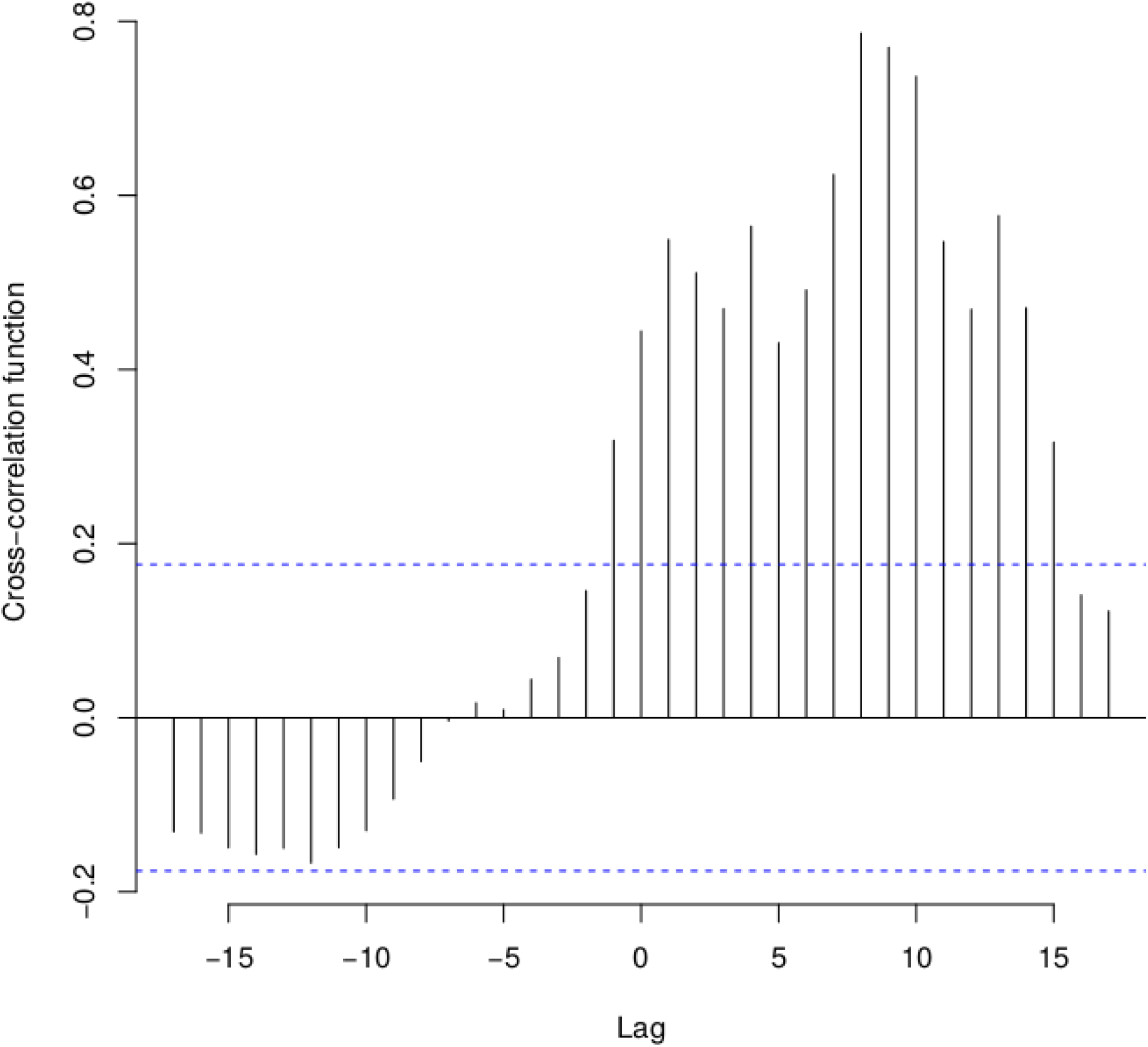
cross-correlation plot of case notifications and SARS-CoV-2 RNA concentration in the wastewater. Negative lags indicate that the RNA concentration is a leading indicator of changes in the RNA concentration.

**Fig. S3:**
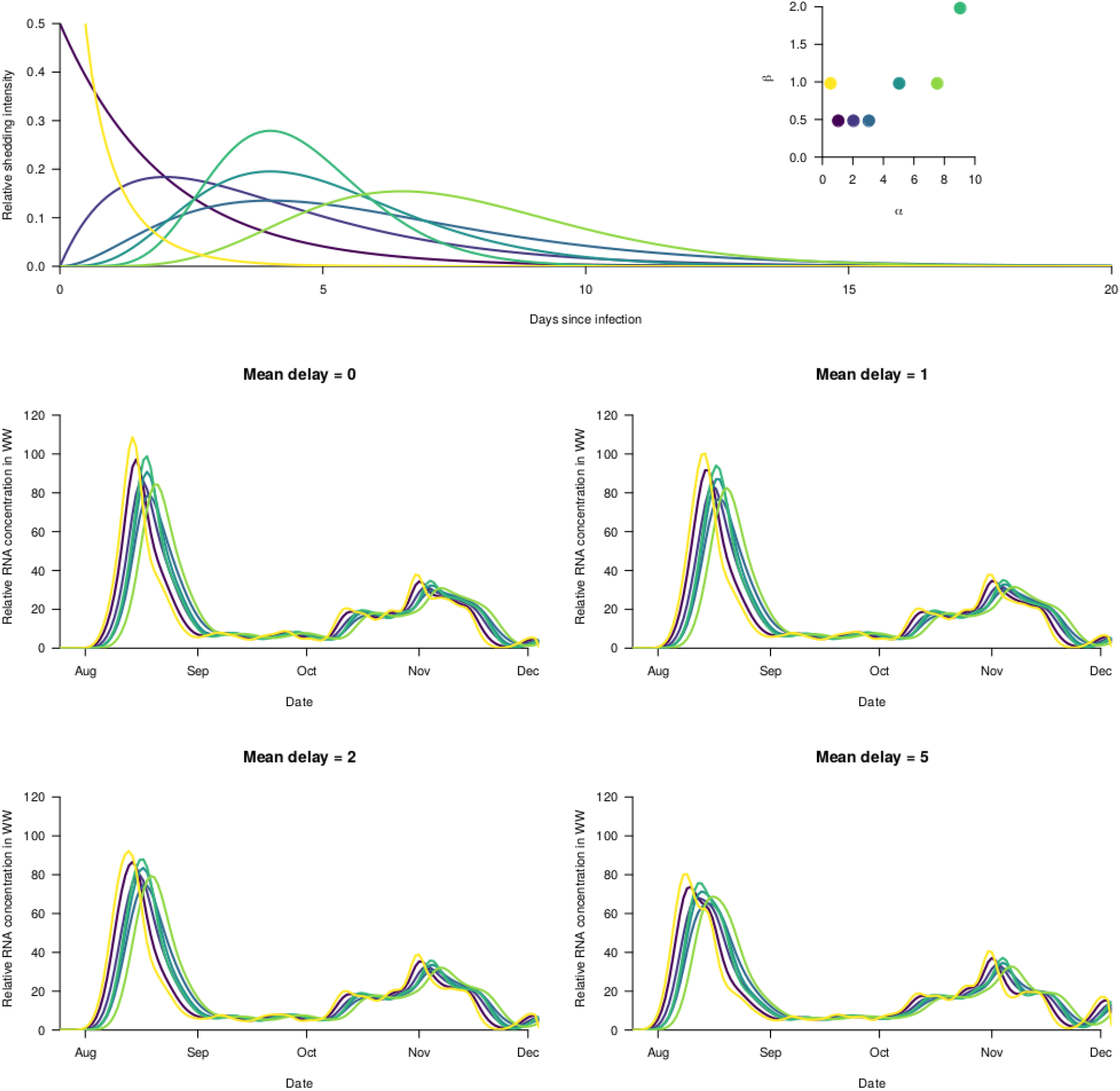
examples of possible shedding distributions. The top panel shows some example (arbitrary) shedding distributions. The bottom four panels show the implied temporal pattern of SARS-CoV-2 in the wastewater for each shedding distribution and for different delays

**Fig. S4:**
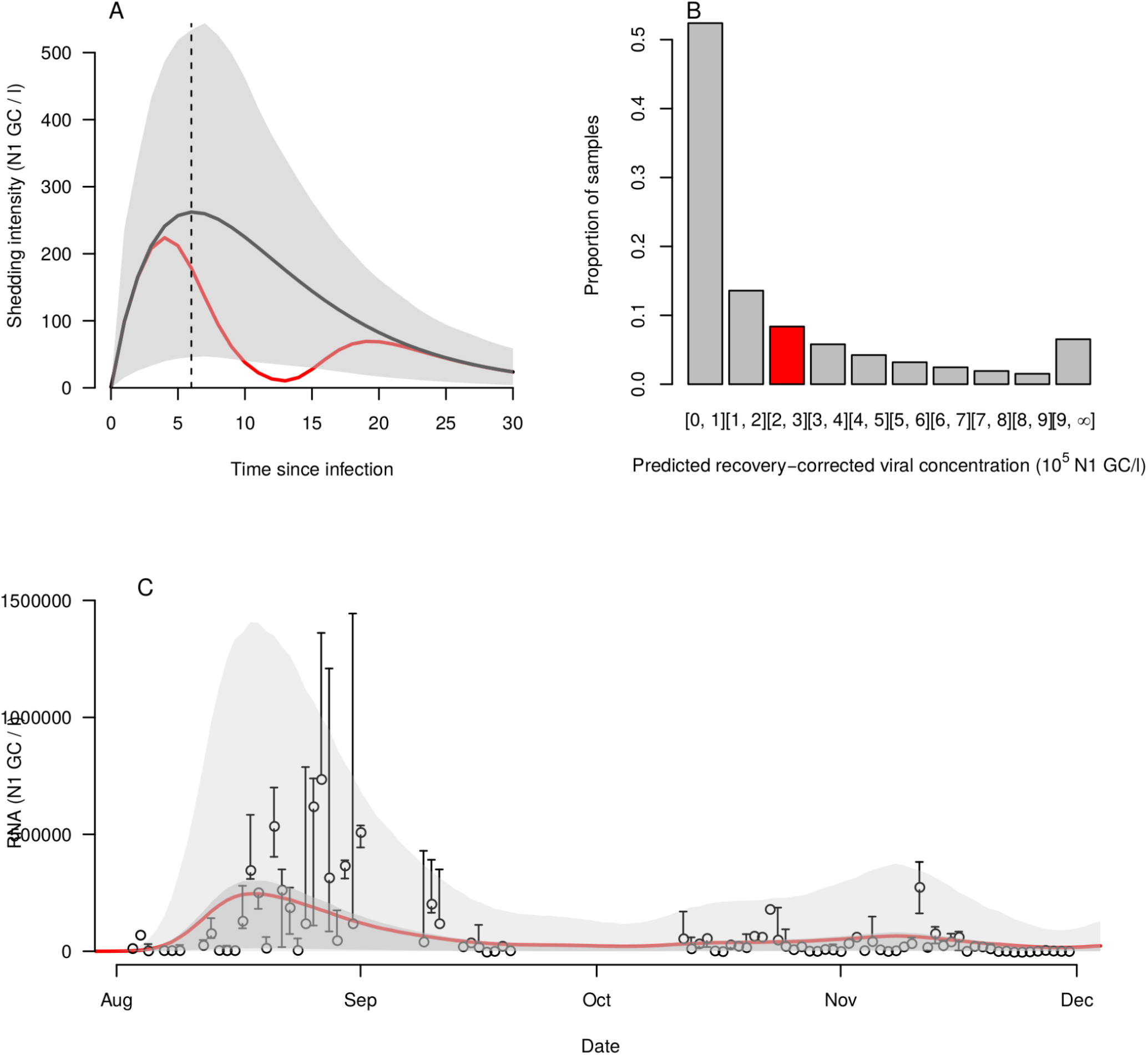
Shedding reconstruction with delay from symptom onset to test of 2, but on a natural rather than a log scale. Otherwise, as in Fig. 2.

**Fig. S5:**
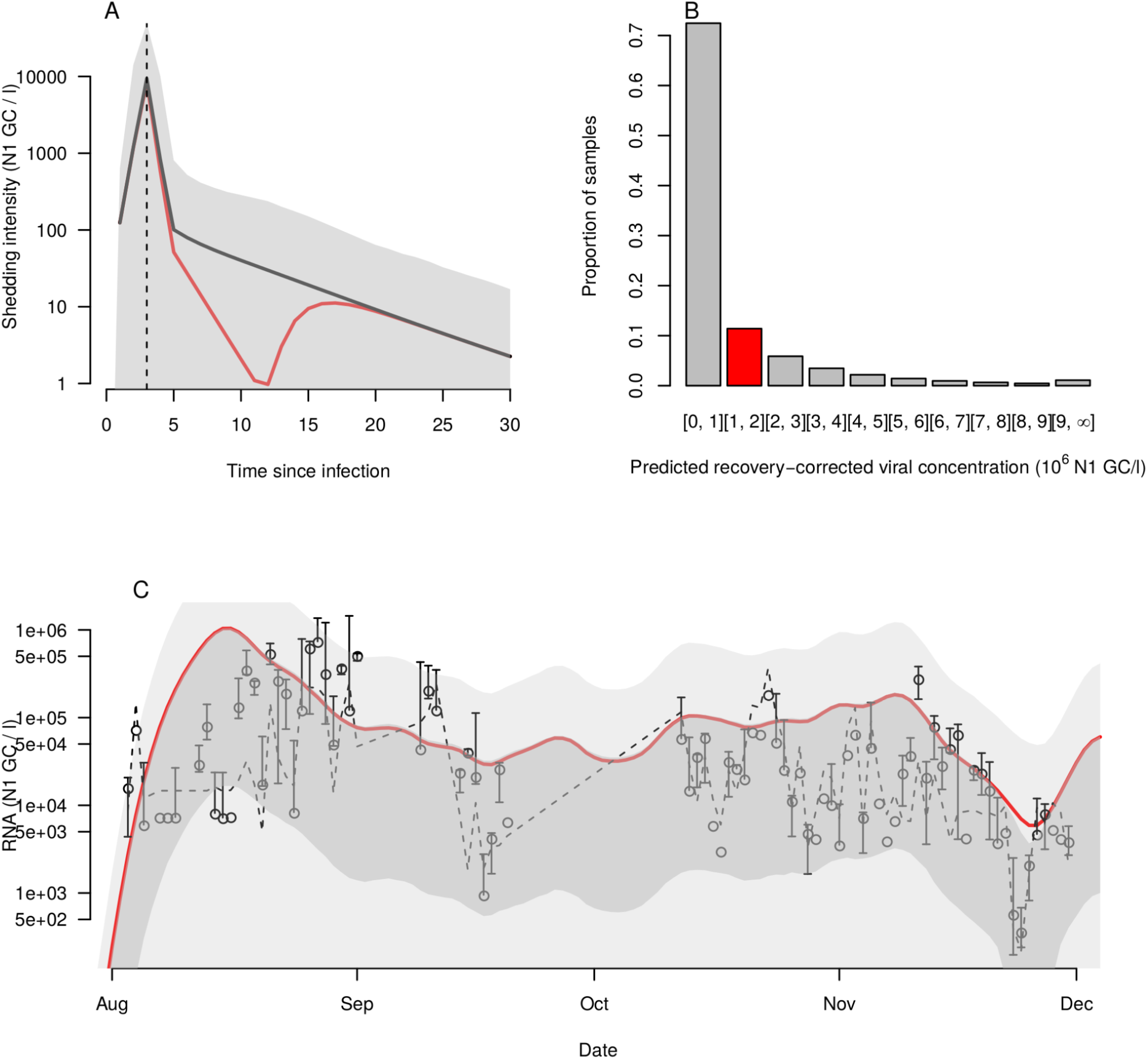
Shedding reconstruction with delay from symptom onset to test of 0. Otherwise, as in Fig. 2.

**Fig. S6:**
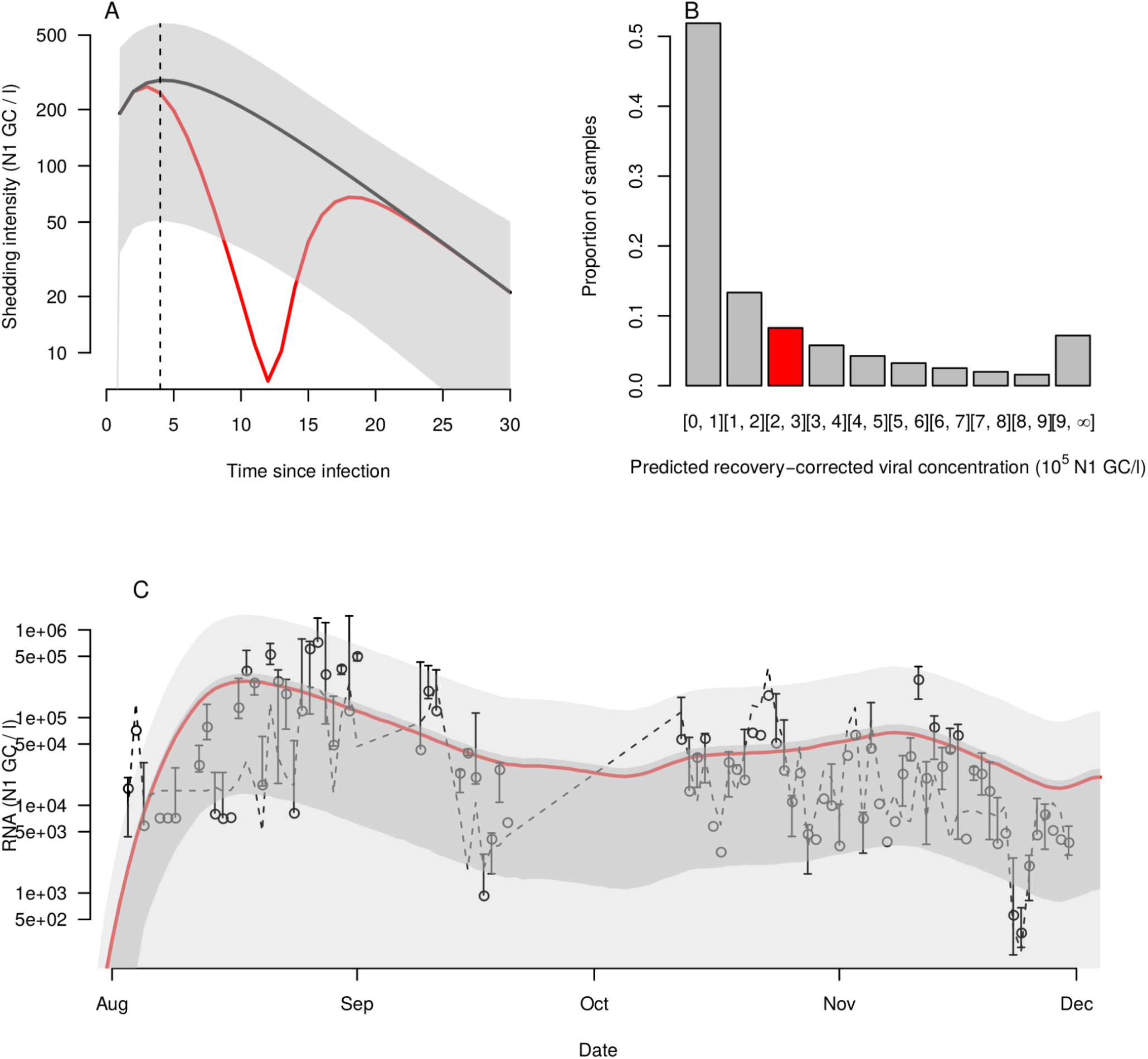
Shedding reconstruction with delay from symptom onset to test of 1 day. Otherwise, as in Fig. 2.

**Fig. S7:**
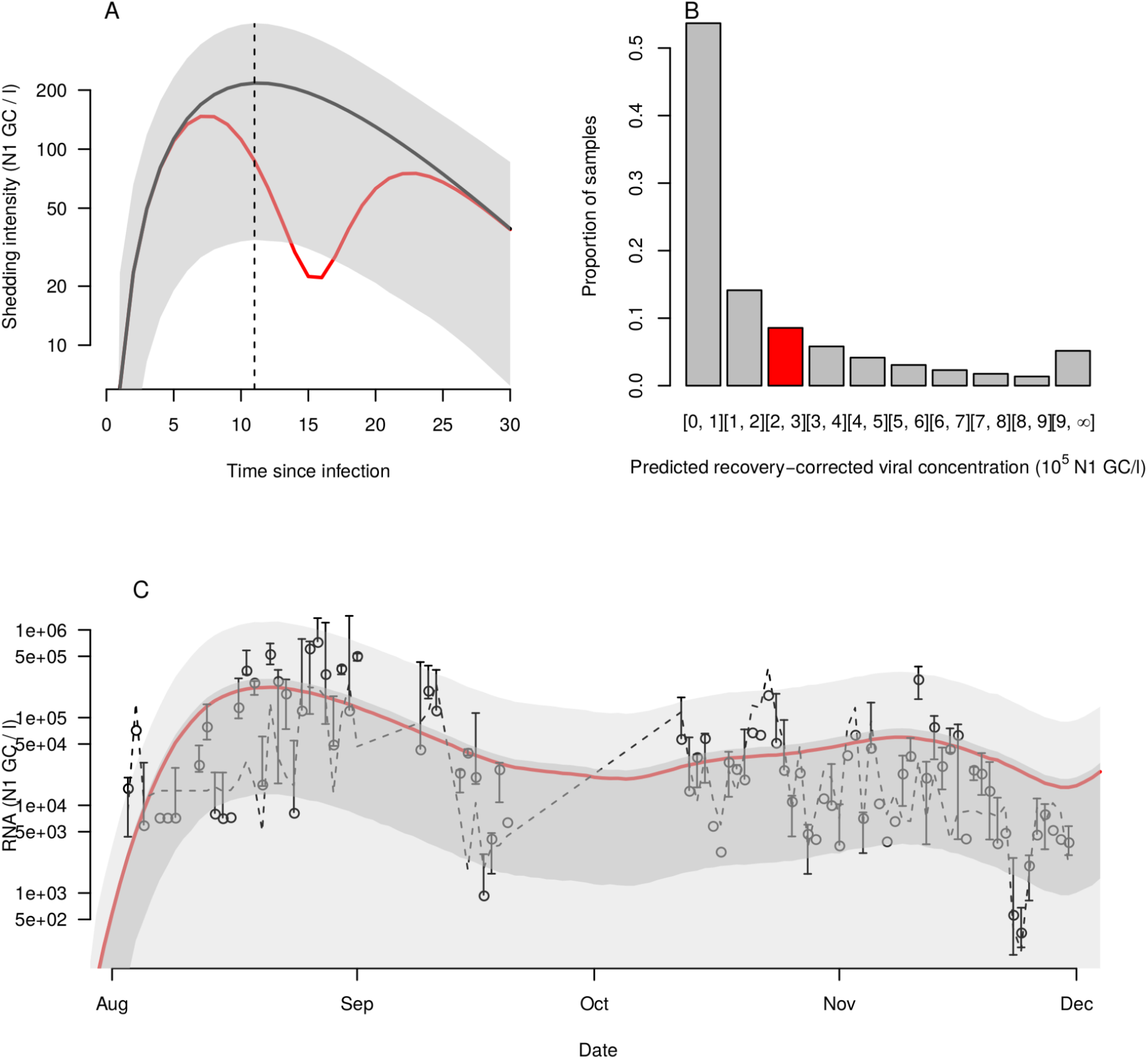
Shedding reconstruction with delay from symptom onset to test of 5 days. Otherwise, as in Fig. 2.

**Fig. S8:**
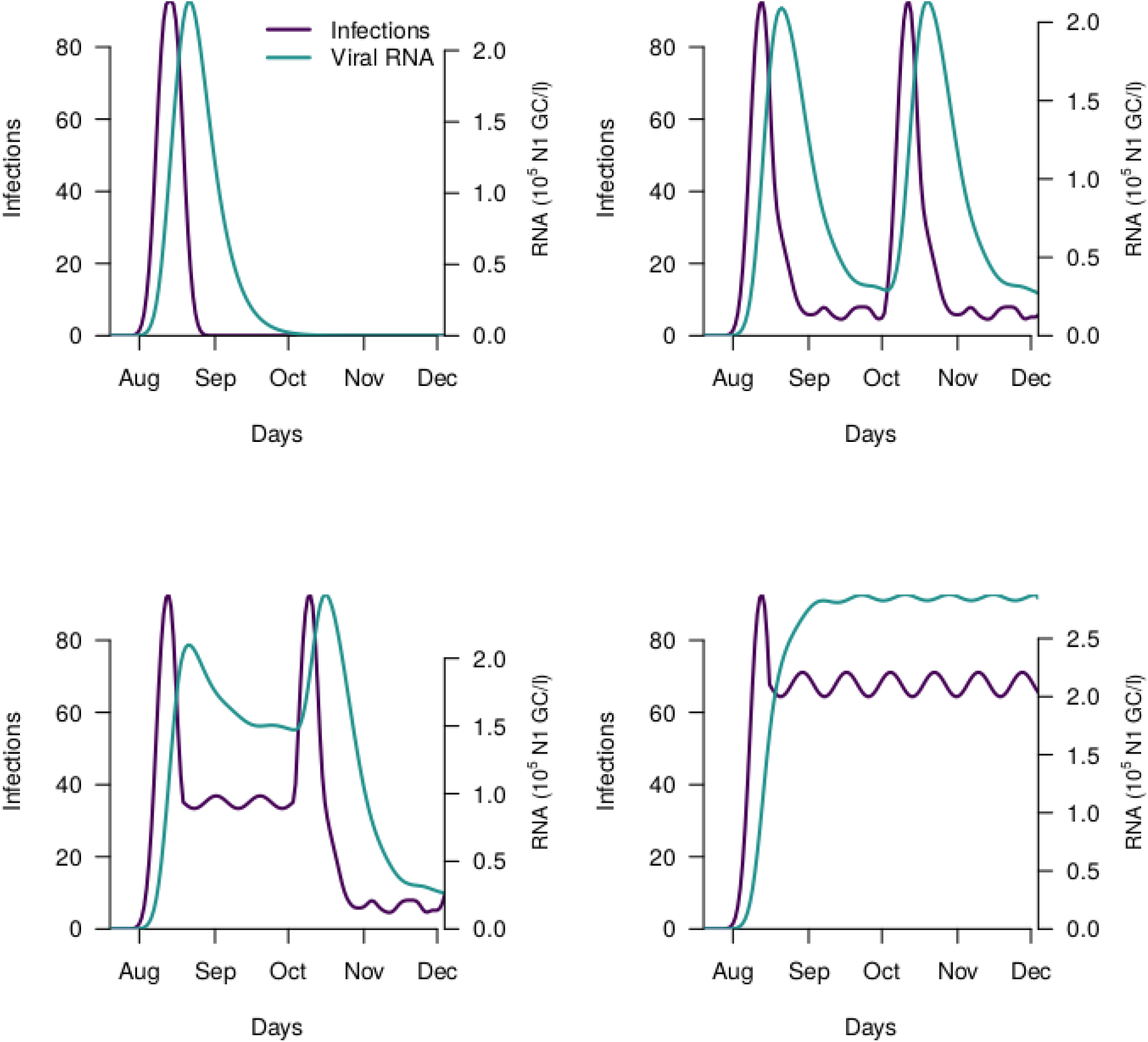
Example mean wastewater concentration timeseries for toy incidence curves, to show the relationship between potential incidence curves and wastewater data. In all panels, a mean delay of two days from symptom onset to test was used. Purple lines show synthetic examplar incidence curves, green lines show the implied mean RNA concentration in wastewater from these synthetic incidence curves.

**Fig. S9:**
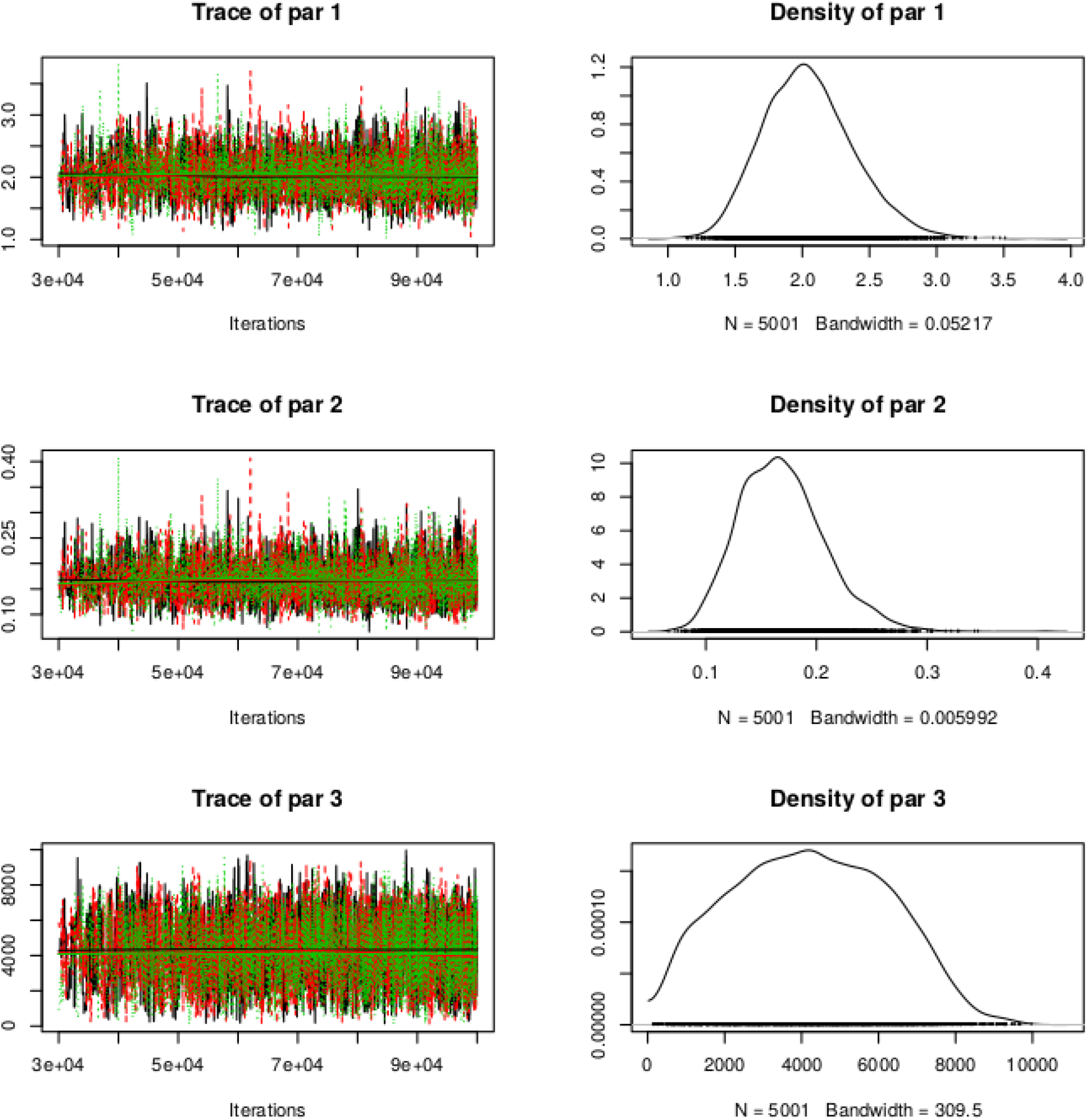
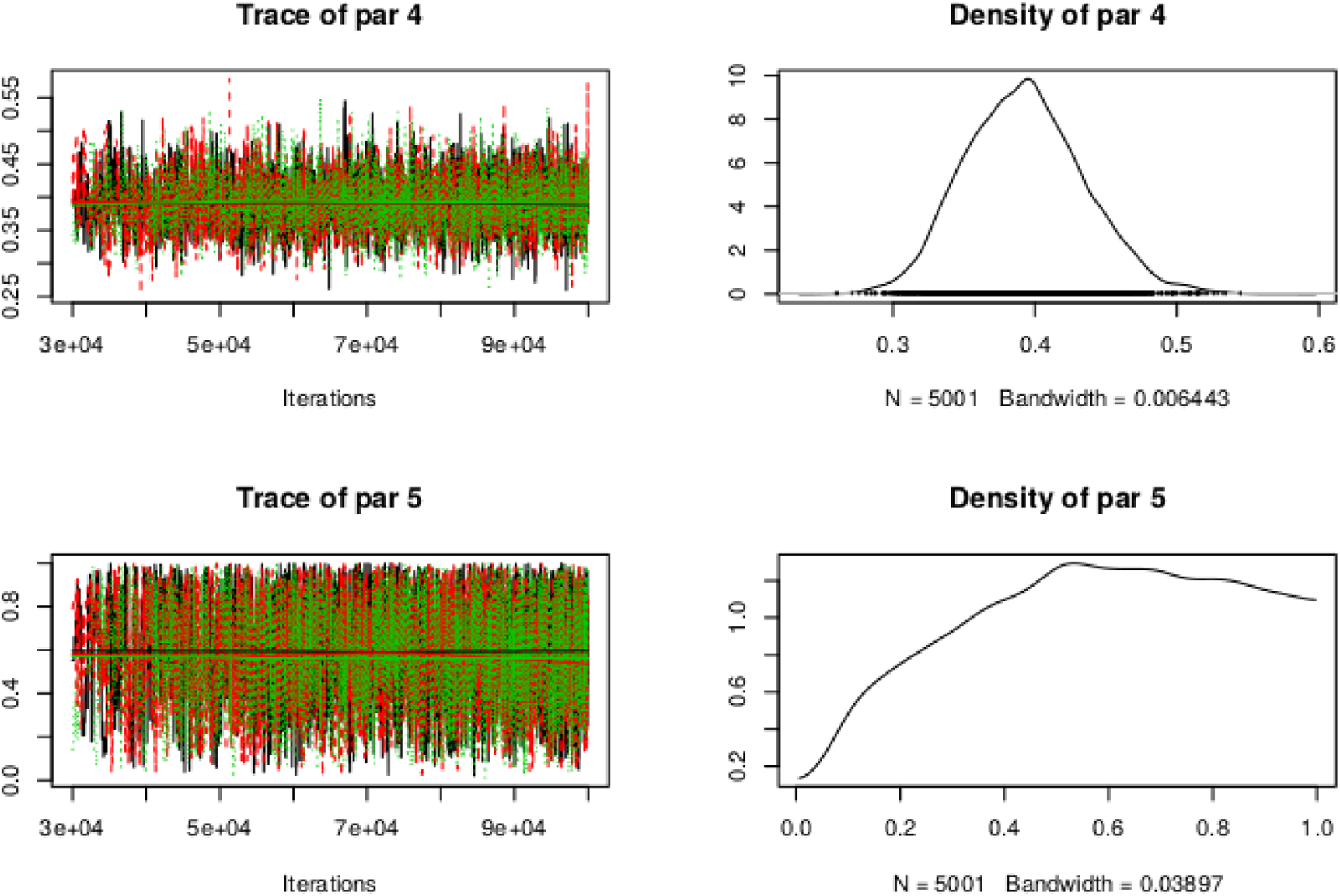
MCMC chains (left column) and posterior parameter densities (right column). par 1 is the gamma shape parameter (α), par 2 is the gamma rate parameter (β), par 3 is the scaling coefficient (θ), par 4 is the negative binomial size parameter (r), and par 5 is the proportion of infections reported (p_r_). There are three chains of 1×10^5^ iterations each, and the first 3×10^4^ iterations of each chain are burned.

**Fig. S10:**
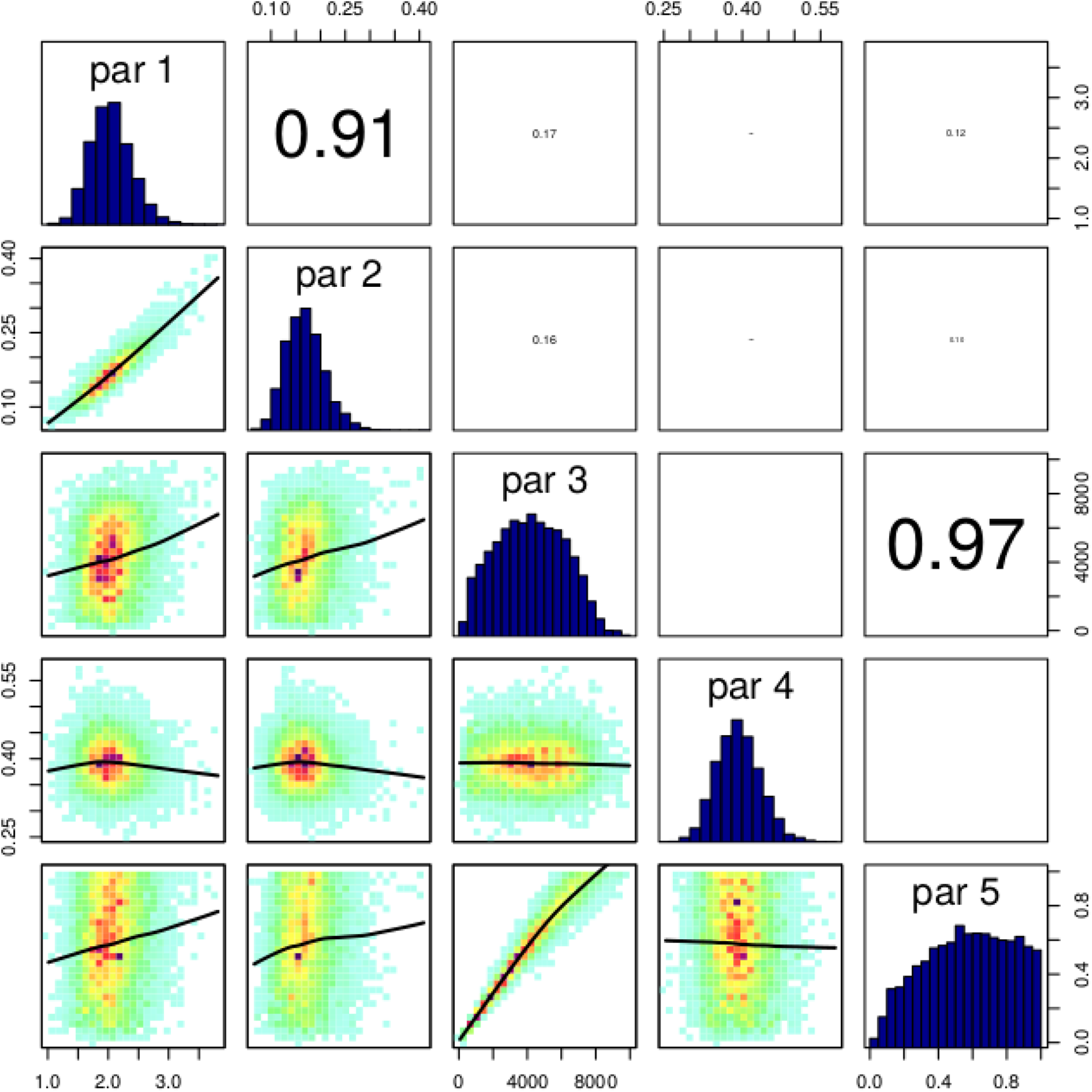
Posterior densities (diagonal), correlation density plots (lower left) and correlation coefficients between parameters (upper right). par 1 is the gamma shape parameter (α), par 2 is the gamma rate parameter (β), par 3 is the scaling coefficient (θ), par 4 is the negative binomial size parameter (r), and par 5 is the proportion of infections reported (p_r_).

## Supplementary table

**Table S1:** Mean values and 95% credible intervals of fitted parameters and DIC for each fitted model. α is the gamma shape parameter, β is the gamma rate parameter, θ is the scaling coefficient, r is the negative binomial size parameter, and p_r_ is the proportion of infections reported.

| Testing delay | 0 days | 1 day | 2 days | 5 days |
| --- | --- | --- | --- | --- |
| $\alpha$ | 460 [1.08 - 980] | 1.6 [1.2 - 2.2] | 2.1 [1.5 - 2.8] | 3.5 [2.5 - 4.7] |
| $\beta$ | 160 [0.0966 - 407] | 0.15 [0.091 - 0.24] | 0.17 [0.10 - 0.25] | 0.23 [0.14 - 0.33] |
| $\theta$ | 8,900 [1,800 - 17,600] | 4,300 [770 - 8,100] | 4,200 [720 - 7,900] | 4,000 [670 - 7,500] |
| $r$ | 0.29 [0.23 - 0.35] | 0.39 [0.31 - 0.47] | 0.39 [0.32 - 0.48] | 0.41 [0.33 - 0.50] |
| $p_r$ | 0.59 [0.18 - 0.98] | 0.57 [0.11 - 0.98] | 0.57 [0.11 - 0.98] | 0.57 [0.10 - 0.97] |
| DIC | 6,400 | 12,000 | 13,000 | 9,700 |

